# Torque teno virus load predicts allograft rejection but not viral infection after kidney transplantation

**DOI:** 10.1101/2021.03.05.21252817

**Authors:** Aline L van Rijn, Herman F Wunderink, Igor A Sidorov, Caroline S de Brouwer, Aloysius CM Kroes, Hein Putter, Aiko PJ de Vries, Joris I Rotmans, Mariet CW Feltkamp

## Abstract

The main challenge of immunosuppressive therapy after solid organ transplantation is to create a new immunological balance that prevents organ rejection and does not promote opportunistic infection. Torque teno virus (TTV) a ubiquitous and non-pathogenic single-stranded DNA virus has been proposed as a marker of functional immunity in immunocompromised patients. Here investigate whether TTV loads predict the risk of common viral infection and allograft rejection in kidney transplantation recipients.

In a retrospective cohort of 389 kidney transplantation recipients, individual TTV loads in were measured by qPCR in consecutive plasma samples during one year follow-up. The endpoints were allograft rejection, BK polyomavirus (BKPyV) viremia and cytomegalovirus (CMV) viremia. Repeated TTV measurements and rejection and infection survival data were analysed in a joint model.

During follow-up, TTV DNA detection in the transplant recipients increased from 85 to 100%. The median viral load increased to 10^7^ genome copies/ml within three months after transplantation. Rejection, BKPyV viremia and CMV viremia occurred in 23%, 27% and 17% of the patients, respectively. With every 10-fold TTV load-increase, the risk of rejection decreased considerably (HR: 0.74, CI 95%: 0.71-0.76), while the risk of BKPyV and CMV viremia remained the same (HR: 1.03, CI 95%: 1.03-1.04 and HR: 1.01, CI 95%: 1.01-1.01).

In conclusion, TTV load kinetics predict allograft rejection in kidney transplantation recipients, but not the BKPyV and CMV infection. The potential use of TTV load levels as a guide for optimal immunosuppressive drug dosage to prevent allograft rejection deserves further validation.

## Introduction

The optimum level of immunosuppression after solid organ transplant (SOTx) varies between individuals and optimal dosing of these essential drugs can be difficult. As a consequence, SOTx recipients experience a number of complications ranging from development of de novo donor specific antibodies and allograft rejection, due to insufficient immunosuppression, to infection as the result of over-immunosuppression. Monitoring individual therapeutic drug levels does not solve this issue, because immunosuppressive trough levels poorly correlate with development of rejection ^1, 2^. Therefore, there is a call for a reliable biomarker of functional immunity in patients that receive immunosuppressive therapy. Such biomarker could provide assistance in balancing the individual immunosuppressive medication and anticipate the risk of both rejection and infection.

Kidney transplantation (KTx) recipients may experience multiple infections of diverse origin. A large proportion is caused by viruses that cause persistent, asymptomatic infection in the general population, but start replicating freely destroying tissues in the absence of functional immunity. BK polyomavirus (BKPyV) for instance, found in blood (viremia) in approximately 30% of KTx recipients, causes BKPyV-associated nephropathy (BKPyVAN) and loss of allograft function in up to 10% ^3^. Furthermore, without prophylaxis cytomegalovirus (CMV) infection occurs in ∼20% of KTx recipients, causing invasive disease involving multiple organs with substantial morbidity and mortality, if left untreated ^4^.

Infection and rejection frequently require adjustment of immunosuppressive medication to compensate for the overimmunosuppression and underimmunosuppression ^3, 5-7^. Unfortunately, a valid biomarker that indicates overimmunosuppression and underimmunosuppression is lacking. Torque teno virus (TTV) load has been proposed as a surrogate marker of functional immunity that might be useful by predicting the risk of rejection and infection in SOTx recipients ^8^.

TTVs are small, single-stranded DNA-viruses that infect everyone without causing documented disease ^9-11^. Thus far twenty-nine species described by the International Committee on Taxonomy of Viruses (ICTV) ^12^. In immune competent individuals, the adaptive cellular immune responses control TTV infection ^13^. TTV blood levels might therefore mirror the efficacy of the immune system in immunosuppressed SOTx patients, with high TTV DNA loads indicating too much immunosuppression and concomitant risk of infection, and low TTV loads indicating too little immunosuppression and risk of rejection.

Several studies have investigated the association between TTV load and infection and rejection in SOTx patients. While some indicated an association between infection and high/increasing TTV load ^14-18^, and some between rejection and low/decreasing TTV load ^14, 15, 19-23^, others were unable to confirm these associations ^24-26^. The ambiguity of these results calls for more systematic study into these associations.

In the present study, we determined TTV load kinetics in blood from KTx recipients drawn before and after KTx, and explored its association with development of kidney rejection and of two common post-KTx viral infections (BKPyV and CMV). A joint model was built to analyse these longitudinal endpoints with the repeated TTV load measurements. With the help of this integrated approach analysing TTV loads against clinical endpoints at opposite ends of the immunosuppression spectrum, the potential use of TTV as a universal biomarker of functional immunity was assessed.

## Materials and Methods

### Cohort and sampling

This study uses a pre-existing retrospective KTx cohort of 407 adult KTx donor and recipient pairs, extensively described by Wunderink et al. ^3, 27, 28^, transplanted between 2003-2013 in the Leiden University Medical Center (LUMC) in the Netherlands. In the current study, only recipients of living donors were included. Blood samples used for TTV DNA detection were collected pre-transplantation, and 1.5, 3, 6, 9 and 12 months after transplantation. If no pre-transplantation sample or less than two post-transplantation samples were available, the recipient was excluded (**Supplement 1**). As a result, 389 KTx recipients with a total of 1663 samples were included. Recorded baseline characteristics are age, sex, underlying renal condition, dialysis vintage and type of maintenance immunosuppressive treatment.

The study protocol was approved by the local scientific committee and submitted to the medical ethics committee of the LUMC, who declared no objection. We adhered to the STROBE statement for reporting observational studies ^29^.

### TTV load (Predictor)

For TTV load detection DNA was extracted from 200 µL of each blood serum and plasma sample, as described previously ^3^. A detailed description of TTV load detection by qPCR can be found in Supplement 2. Measured TTV loads were log_*10*_-transformed, as per general convention for viral load. Determined TTV loads below the LOD were set to LOD/√2 to approximate the assumed normal distribution of very weakly positive and negative loads ^33^.

### BKPyV viremia and CMV viremia (Infection endpoints)

The first and second endpoint of this study were the development of BKPyV and CMV viremia, respectively, defined as the first day after transplantation on which viral DNA is detected in the blood. The presence of BKPyV DNA in the samples was determined by qPCR in a previous study ^3^. CMV load data were obtained from blood plasma samples previously collected based on clinical suspicion and analysed by qPCR for the presence of CMV DNA ^34^. The median follow-up was 9.36 months for BKPyV viremia and 12 months for CMV viremia, calculated with the reverse Kaplan Meier method ^35^, because participants without BKPyV viremia were censored on the day of their last BKPyV screening. From 2008 onwards, all recipients received CMV prophylaxis for 90 days except for seronegative recipients with a seronegative donor. (**Table 2**).

### Kidney allograft rejection (Rejection endpoint)

Allograft rejection was the third endpoint in this study. Allograft rejection was defined as the first initiation of rejection treatment after transplantation. In some patients, rejection treatment was initiated without prior histological confirmation of allograft rejection if clinical suspicion was high and alternative explanations were excluded. Suspicion of rejection included increased serum creatinine levels, low concentration of immunosuppressive medications and allograft biopsy with histological evidence of rejection. First rejection treatment consisted of 1000 mg methylprednisolone intravenously for three days. The median follow-up for rejection was 12 months, calculated with the reverse Kaplan Meier method ^35^, meaning that every participant completed the full follow-up period if they did not develop rejection.

### Statistical analysis

Statistical software R version 3.5.3 was used for all statistics ^36^. The figures, survival analyses and joint models were made with the appropriate R packages ^37-41^. The baseline characteristics were compared with chi-squared or two-tailed Student’s t-test.

A linear mixed effects model was fitted on the TTV loads. This model models the mean progression of the TTV load over time, using effects that are the same for every individual – the fixed effects – and effects that are unique for every individual – the random effects. A detailed description of the linear mixed effects model can be found in **Supplement 3**. The model was combined with a survival analysis for the endpoints, in a so-called joint model analysis ^40^. This model estimates the risk rate of the events in cox-proportional hazards model, based on the modelled TTV loads. Three joint model analyses were performed, to accommodate the survival analysis for each endpoint - BKPyV viremia, CMV viremia and rejection. The association between a 1 log change in TTV load and the time-to-event is reported as hazard ratio (HR).

## Results

### Incidence of viremia, and relation to population baseline characteristics

During follow-up, 105 out of 389 KTx recipients (27%) developed BKPyV viremia and 77 (20%) developed CMV viremia within one year after transplantation (**Table 1, Figure 1**). Baseline characteristics were comparable across BKPyV viremia and non-BKPyV viremia groups. CMV viremia was observed less often after 2008 (**Table 2**), which could be related to the start of CMV prophylaxis use ^42^. Also, in 2008, the use tacrolimus was initiated (11% before, 97% during and after 2008). This explains why the non-CMV viremia group had received tacrolimus more often (**Table 1**).

**Table 1:**
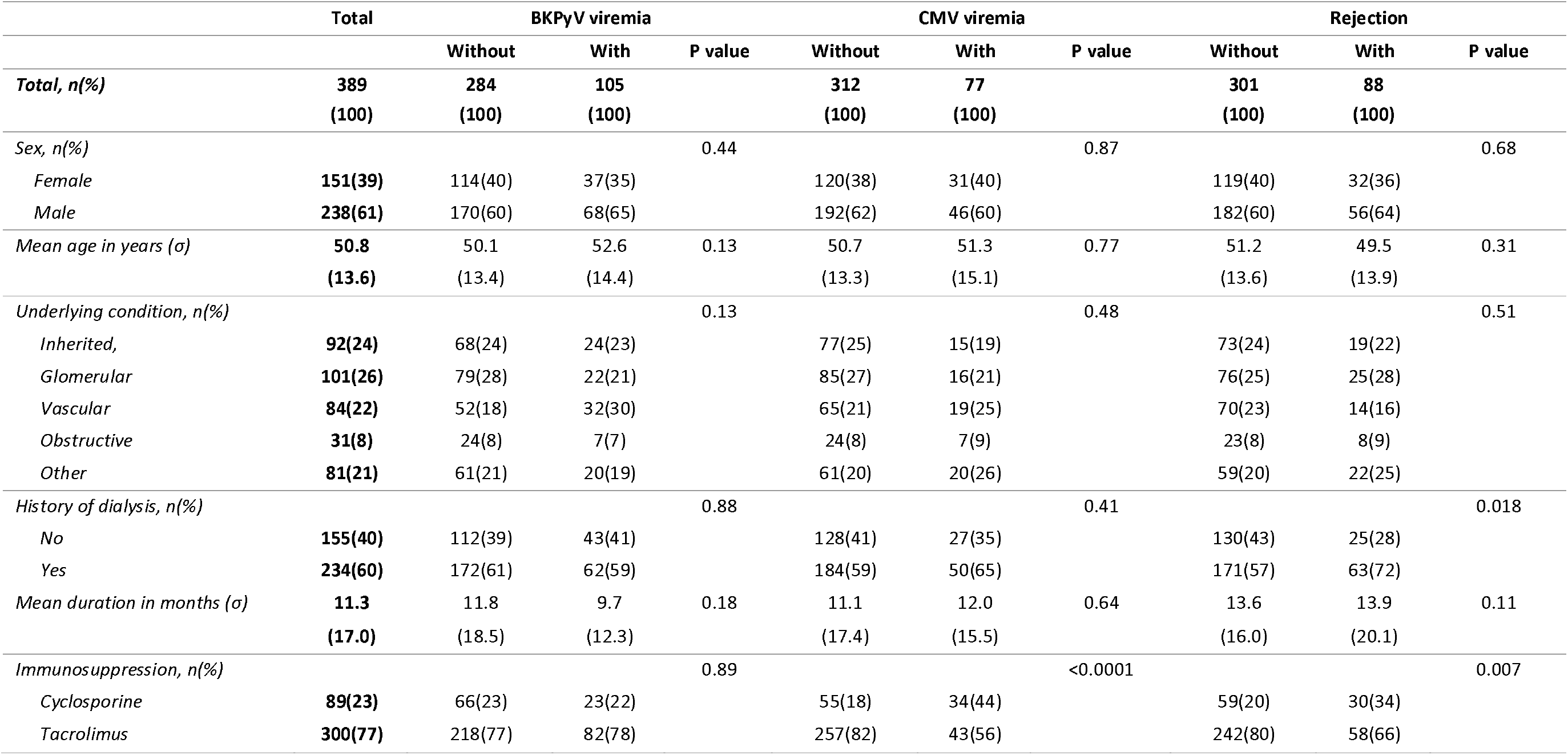
Baseline characteristics of the study population

|  | Total | BKPyV viremia |  |  | CMV viremia |  |  | Rejection |  |  |
| --- | --- | --- | --- | --- | --- | --- | --- | --- | --- | --- |
|  |  | Without | With | P value | Without | With | P value | Without | With | P value |
| **Total, n(%)** | **389**<br>**(100)** | **284**<br>**(100)** | **105**<br>**(100)** |  | **312**<br>**(100)** | **77**<br>**(100)** |  | **301**<br>**(100)** | **88**<br>**(100)** |  |
| *Sex, n(%)* |  |  |  | 0.44 |  |  | 0.87 |  |  | 0.68 |
| Female | 151(39) | 114(40) | 37(35) |  | 120(38) | 31(40) |  | 119(40) | 32(36) |  |
| Male | 238(61) | 170(60) | 68(65) |  | 192(62) | 46(60) |  | 182(60) | 56(64) |  |
| *Mean age in years ($\sigma$)* | **50.8**<br>**(13.6)** | 50.1<br>(13.4) | 52.6<br>(14.4) | 0.13 | 50.7<br>(13.3) | 51.3<br>(15.1) | 0.77 | 51.2<br>(13.6) | 49.5<br>(13.9) | 0.31 |
| *Underlying condition, n(%)* |  |  |  | 0.13 |  |  | 0.48 |  |  | 0.51 |
| Inherited, | 92(24) | 68(24) | 24(23) |  | 77(25) | 15(19) |  | 73(24) | 19(22) |  |
| Glomerular | 101(26) | 79(28) | 22(21) |  | 85(27) | 16(21) |  | 76(25) | 25(28) |  |
| Vascular | 84(22) | 52(18) | 32(30) |  | 65(21) | 19(25) |  | 70(23) | 14(16) |  |
| Obstructive | 31(8) | 24(8) | 7(7) |  | 24(8) | 7(9) |  | 23(8) | 8(9) |  |
| Other | 81(21) | 61(21) | 20(19) |  | 61(20) | 20(26) |  | 59(20) | 22(25) |  |
| *History of dialysis, n(%)* |  |  |  | 0.88 |  |  | 0.41 |  |  | 0.018 |
| No | 155(40) | 112(39) | 43(41) |  | 128(41) | 27(35) |  | 130(43) | 25(28) |  |
| Yes | 234(60) | 172(61) | 62(59) |  | 184(59) | 50(65) |  | 171(57) | 63(72) |  |
| *Mean duration in months ($\sigma$)* | **11.3**<br>**(17.0)** | 11.8<br>(18.5) | 9.7<br>(12.3) | 0.18 | 11.1<br>(17.4) | 12.0<br>(15.5) | 0.64 | 13.6<br>(16.0) | 13.9<br>(20.1) | 0.11 |
| *Immunosuppression, n(%)* |  |  |  | 0.89 |  |  | <0.0001 |  |  | 0.007 |
| Cyclosporine | 89(23) | 66(23) | 23(22) |  | 55(18) | 34(44) |  | 59(20) | 30(34) |  |
| Tacrolimus | 300(77) | 218(77) | 82(78) |  | 257(82) | 43(56) |  | 242(80) | 58(66) |  |

**Table 2:** CMV serostatus of donor and recipient.

| CMV<br>serostatus | Total | Before 2008 |  | After 2008 <sup>a</sup> |  |
| --- | --- | --- | --- | --- | --- |
|  |  | No CMV<br>viremia | CMV<br>viremia | No CMV<br>viremia | CMV<br>viremia |
|  | **389** | **59 (66%)** | **31 (34%)** | **265 (89%)** | **34 (11%)** |
| R-/D- | **86** | 20 (100%) | 0 (0%) | 66 (100%) | 0 (0%) |
| R+/D+ | **155** | 25 (64%) | 14 (36%) | 105 (91%) | 11 (9%) |
| R-/D+ | **84** | 11 (52%) | 10 (48%) | 42 (67%) | 21 (33%) |
| R+/D- | **64** | 3 (30%) | 7 (70%) | 52 (96%) | 2 (4%) |

R: recipient, D: donor. <sup>a</sup>: After 2008, all except R-/D- received valganciclovir CMV prophylaxis for 90 days after transplantation.

**Figure 1:**
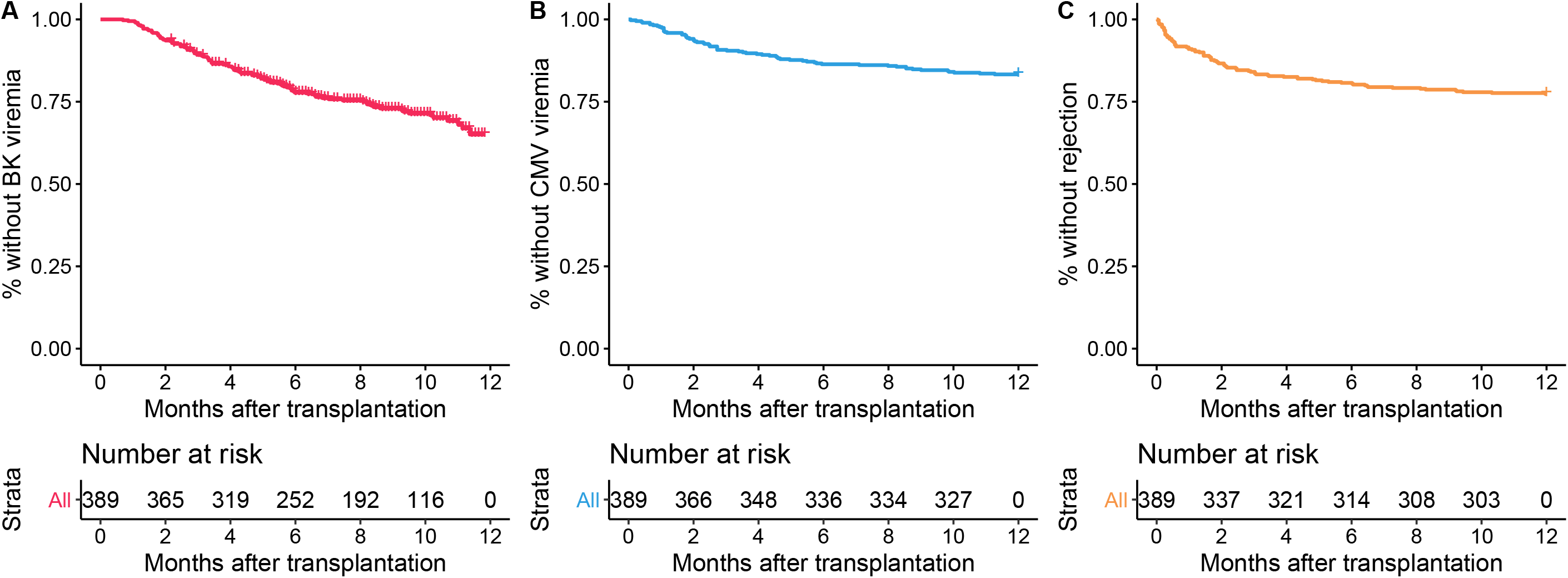
Development of BKPyV viremia, CMV viremia and allograft rejection after KTx. Survival curves are shown for BKPyV viremia (A), CMV viremia (B), and allograft rejection (C) after KTx.

### Incidence of rejection, and relation to population baseline characteristics

Allograft rejection developed in 88 KTx recipients (23%) within one year after KTx (**Figure 1**). Nineteen percent (58/300) of patients who received tacrolimus developed rejection, opposed to 34% (30/89) of patients receiving cyclosporine A, which is probably related to the larger immunosuppressive potential of tacrolimus over cyclosporine A (**Table 1**) ^43^. Furthermore, 72% (63/88) of the recipients with rejection had a history of dialysis, compared to 57% (171/301) of the recipients without rejection. This association was also found in the joint model analysis discussed below. This association might have been confounded by a lower degree of HLA mismatch, since pre-emptive transplantations often involve family members.

### TTV load kinetics

TTV DNA load was determined in 1663 samples from 389 eligible patients, with 2-5 measurements per patient. A visualisation of the measured TTV loads over time is shown in **Figure 2**. The median TTV load at baseline was the equivalent of 5012 genome copies/ml which is 3.7 log (Inter quartile range (IQR) 2.6-4.7). TTV loads below the LOD were observed in 15% of the subjects. During one year follow-up after transplantation, detectable TTV loads were obtained in all recipients. The median TTV load detected was 5.2 log copies/ml (IQR 3.9-6.5) at 1.5 months, 7.4 (IQR 5.9-8.9) at 3 months, 6.1 (IQR 4.5-7.8) at 6 months, and 5.2 (IQR 4.0-6.4) at 12 months.

**Figure 2:**
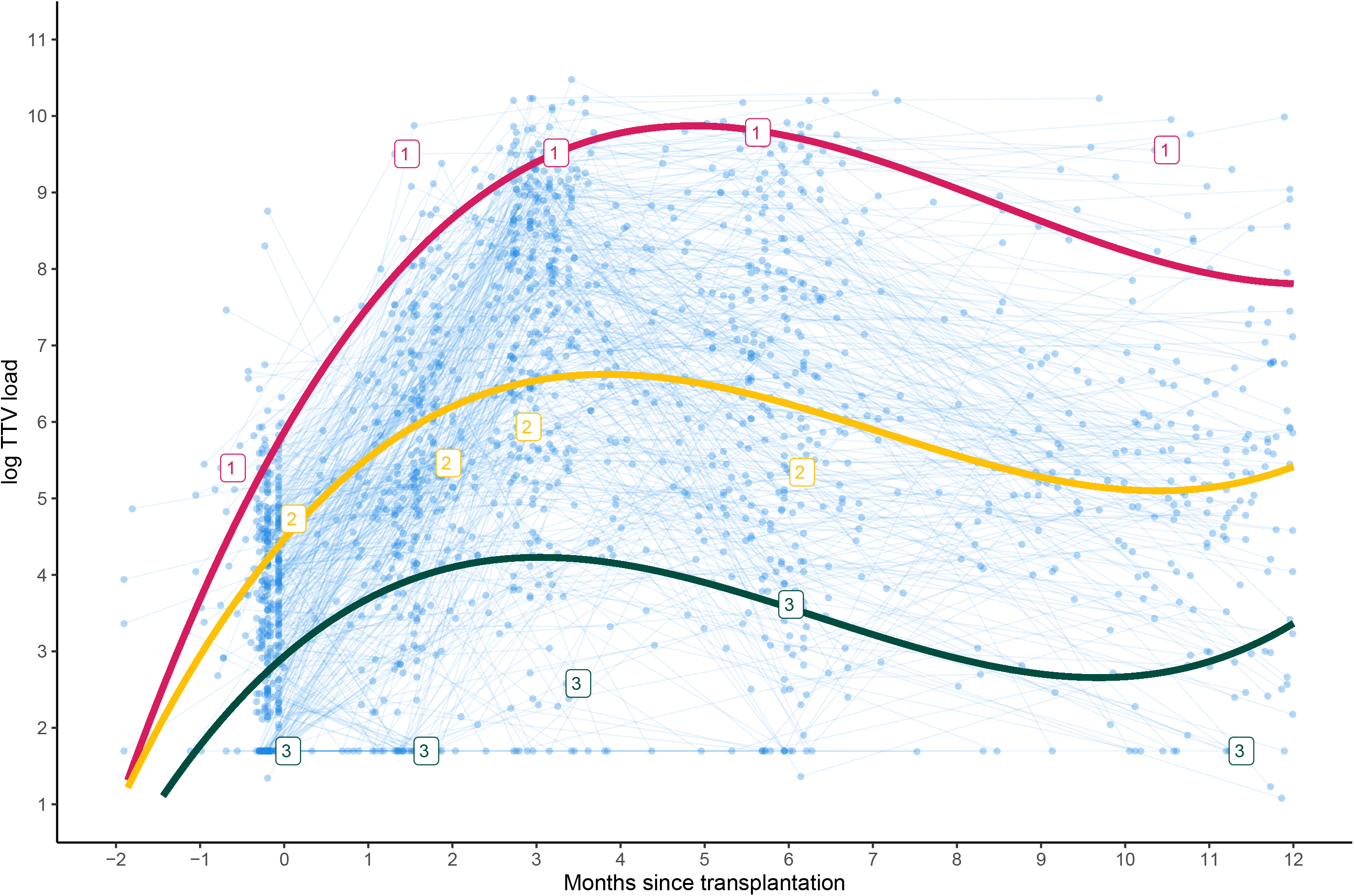
Observed and modelled TTV loads after KTx. Individual TTV DNA load measurements are depicted in blue, shown for each individual sample by the dots connected by a line. The red, orange and blue labels and lines depict the observed and TTV loads predicted by the linear mixed model of three example patients. Taking into account their individual random effects, the individual linear progression trajectories can be predicted with the formula described in Materials and Methods, the coefficient values of the fixed effects in Table 3, and their unique random effects: Patient 1 (red): tacrolimus = yes, dialysis vintage = yes, βi0 = 1.58, βi1 = 0.43, βi2 = 0.03 Patient 2 (yellow): tacrolimus = no, dialysis vintage = yes, βi0 = 0.18, βi1 = 0.19, βi2 = 0.01 Patient 3 (green): tacrolimus = yes, dialysis vintage = no, βi0 = 1.35, βi1 = 0.52, βi2 = 0.04

### TTV load model

For further analysis, the TTV loads were modelled in a linear mixed effects model (**Table 3**). The final model contained the baseline TTV load (*β*_0_), and fixed coefficients, *β*_1_, *β*_2_,, *β*_3_, *β*_4_ and, *β*_5_, written in formula as: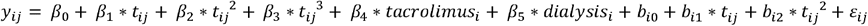 The other tested covariates (sex, age, and underlying condition; **Table 1**) did not improve the prediction of the progression of the TTV load over time. Receiving tacrolimus instead of cyclosporine A and having a history of dialysis both correspond to a higher TTV load (β_4_: 0.51, 95% confidence interval (CI): 0.16-0.86, β_5_: 0.46, 95% CI:0.16-0.76). The variation in the random effects per individual, *b*_*i*0_, *b*_*i*1_ and *b*_*i*2_, and the residual variation over all individuals and time points, *ε*_*ij*_, is reported as standard deviation (σ) around the mean (**Table 3**). To illustrate the individualized predictions of the linear mixed model, three example patients with their observed and predicted TTV loads are shown in **Figure 2**.

**Table 3:** Fixed and random effects from the linear mixed effects model for estimating the mean TTV load over time (t)

| Fixed Effects | β-coefficients | 95% CI | | P value | Random effects | $\sigma$ |
| --- | --- | --- | --- | --- | --- | --- |
| $\beta_0$ *Intercept* | 3.39 | 3.07 | 3.70 | <0.001 | $b_{i0}$ *Intercept* | 0.87 |
| $\beta_1$ $t$ | 1.471 | 1.34 | 1.60 | <0.001 | $b_{i1}$ $t$ | 0.52 |
| $\beta_2$ $t^2$ | -0.24 | -0.27 | -0.21 | <0.001 | $b_{i2}$ $t^2$ | 0.04 |
| $\beta_3$ $t^3$ | 0.01 | 0.01 | 0.01 | <0.001 | $\varepsilon_{ij}$ *Residual* | 1.43 |
| $\beta_4$ *Tacrolimus vs. Cyclosporine A* | 0.45 | 0.15 | 0.74 | 0.0034 | | |
| $\beta_5$ *History of dialysis* | 0.36 | 0.10 | 0.60 | 0.0062 | | |

### TTV load and development of infection and rejection

To compare the TTV load kinetics with the study endpoints, three joint models were built; for BKPyV viremia, CMV viremia and rejection, respectively. The joint model analyses for BKPyV and CMV showed no association between changing TTV loads and development of viremia, with hazard ratios close to one (1.03 95% CI: 1.03-1.04 and 1.01 95% CI: 1.00-1.01 respectively; **Table 4**). For rejection a significant, inverse association was found between increasing TTV load and the time-to-rejection: with every 10-fold (1 log) TTV load-increase, the risk of rejection decreased with a HR of 0.74 (95% CI: 0.71-0.76) (**Table 4**).

**Table 4:** Estimated hazard rate for BKPyV viremia and rejection based on changes in TTV load after KTx

|  |  | **HR\*** | **95% CI** |  | **P value** |
| --- | --- | --- | --- | --- | --- |
| **BKPyV viremia** | *TTV load 1 log increase* | 1.03 | 1.03 | 1.04 | <0.001 |
| **CMV viremia** | *TTV load 1 log increase* | 1.01 | 1.01 | 1.01 | <0.001 |
| **Rejection episode** | *TTV load 1 log increase* | 0.74 | 0.71 | 0.76 | <0.001 |

\* HR: hazard rate

## Discussion

With the help of joint modelling of KTx cohort data, we analysed TTV load kinetics after KTx and explored the association between TTV load and frequent complications of immunosuppression. We showed that an increase of TTV load corresponds to a lower risk of rejection and an equal risk of CMV and BKPyV infection. This observation partially supports the hypothesis that the TTV load reflects the functional immune status of KTx patients. Furthermore, it puts TTV forward as a useful biomarker of functional immunity in SOTx patients, to predict allograft rejection.

The observed effect of TTV load on the risk of developing rejection was quite strong (HR of 0.74 per log TTV load increase), especially if one considers the median TTV load-increase of 3 log observed after transplantation. This implies that patients with substantial TTV load increase, like Patient 1 in Figure 2, have a much lower chance of developing rejection than Patients 2 and 3 who displayed a limited TTV load increase. More detailed study is needed to pinpoint the period after KTx that offers the biggest power to predict the chance of rejection.

Our results are in line with findings from earlier, smaller studies that reported an inverse association between TTV load and rejection ^23, 44^, and the absence of association between TTV load and BKPyV and CMV infection ^26^. However, some studies did show an association between TTV load-increase and infectious complications. This discrepancy may be due to differences in study population, start and duration of follow-up, pathogens studied and/or modelling method ^45-47^.

Our observation that the risk of BKPyV viremia was not related with TTV-load nor with any of the other covariates, fits with findings from one of our previous studies aimed at identifying risk factors for BKPyV infection ^3^. In that study performed within the same cohort, we showed that the risk of BKPyV viremia is primarily governed by the BKPyV infection risk imposed by the donor, which can be estimated based on pre-KTx donor BKPyV IgG seroreactivity, and much less dependent on the immune status of the recipient.

Comparable to BKPyV, we did not find an association between the incidence of CMV viremia and TTV load. However, in the case of CMV, the lack of such an association is more difficult to interpret, since it could be partially masked by the use of anti-viral prophylaxis (**Table 2**). Prophylaxis-stratified analyses were attempted but the model failed to converge due to insufficient observations ^48, 49^. Additional stratified analyses with larger sample sizes are advisable for future research.

When analysing the TTV load kinetics with the help of a mixed model, it is clear that the variation in random effects largely explains the interpatient variability in TTV load. This is because the variability is not explained by the covariates we tested, except for type of immune suppression and dialysis vintage. This promotes the potential value of TTV as a biomarker even more, because interpatient variability in TTV loads signifies high-risk and low-risk patients on top of the known risk factors.

Our study has several strengths that increase the validity of the findings. First of all, the used PCR primer set targets several TTV species at once, which is favourable since a recent study shows the interindividual variability in TTV species is large ^50^. Second, the use of a joint model allows the longitudinal TTV measurements to be used as predictor of time to viremia and to rejection. A common pitfall of analysing this type of data is introducing ‘immortal time’ bias, which is circumvented by the joint model approach.

Regarding potential weaknesses, our study might have overestimated the actual number of rejection episodes. Of 88 patients with high clinical suspicion of a rejection episode, 80 had undergone a biopsy, and in 68% of them there was clear histological evidence for rejection, which may cause an underestimation of the true effect. Future study with biopsy-confirmed rejection is needed for a more precise approximation of the predictive effect of the TTV load.

In conclusion, while TTV load changes fail to predict the risk of BKPyV and CMV infection within the first year after KTx, they strongly predict the development of allograft rejection. Hence, the use of TTV loads as a predictive biomarker for allograft rejection and optimal immunosuppressive dosage deserves further exploration, in KTx patients and in SOTx patients in general, as it could improve patient care and allograft survival.

## Data Availability

The data that support the findings of this study are available on request from the corresponding author, AvR. The data are not publicly available due to privacy and ethical reasons.

## Abbreviations

BKPyV: BKpolyomavirus
CI: Confidence interval
CNI: Calcineurin inhibitor
ESDR: End stage renal disease
HR: Hazard ratio
IQR: Inter quartile range
KTx: Kidney transplantation
LOD: Limit of detection
LUMC: Leiden University Medical Center
PCR: Quantitative PCR
SOTx: Solid organ transplantation
t: time
TTV: Torque teno virus

## Description of supporting information

**Supplement 1:** Flowchart for inclusion and exclusion of kidney transplantation recipients.

**Supplement 2:** A detailed description of TTV detection

**Supplement 3:** A detailed description of the linear mixed effects model

---

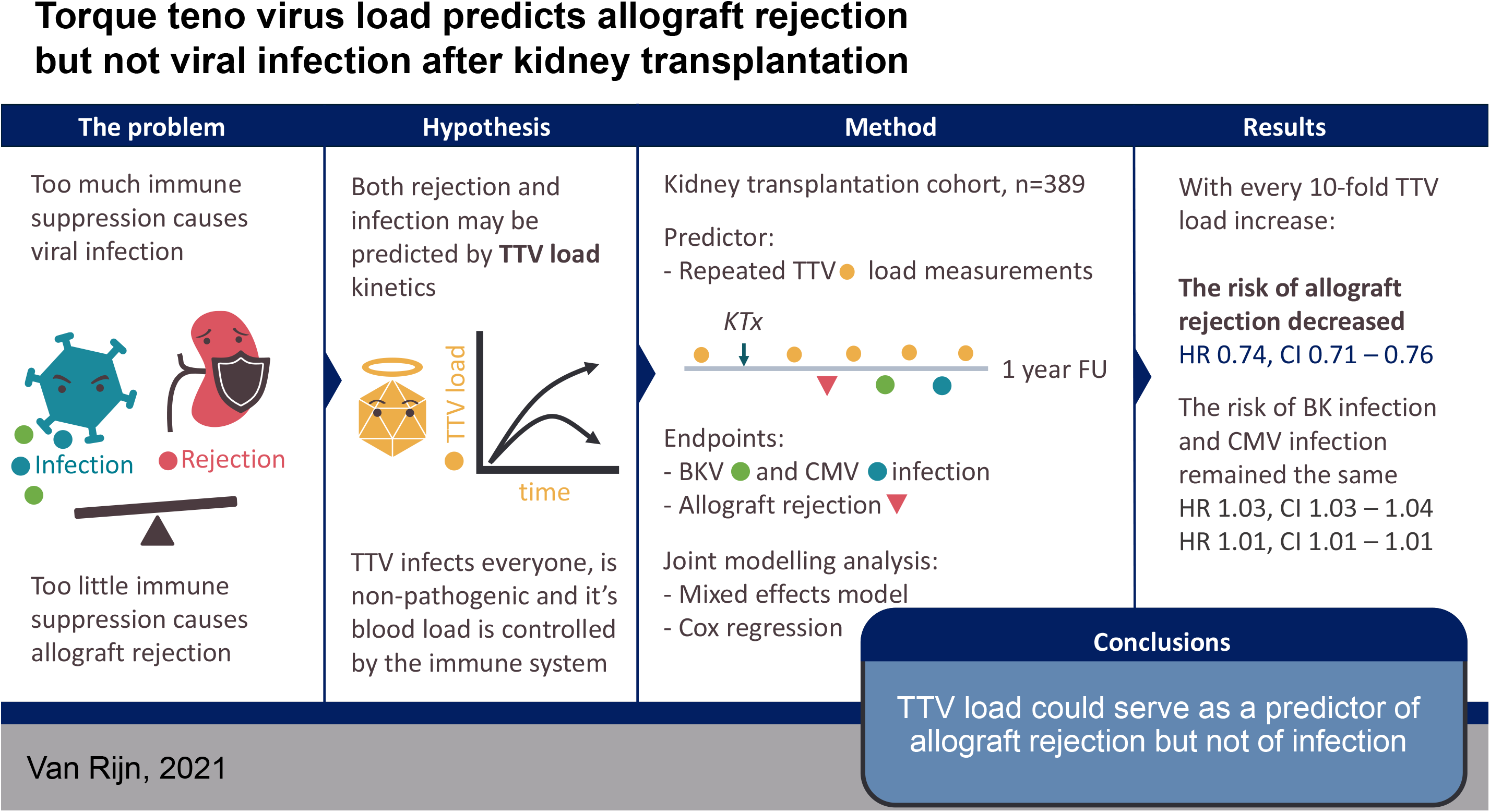

## References

1. Bouamar R, Shuker N, Hesselink DA, et al. Tacrolimus predose concentrations do not predict the risk of acute rejection after renal transplantation: a pooled analysis from three randomized-controlled clinical trials(dagger). Am J Transplant. May 2013;13(5):1253–61. doi:10.1111/ajt.12191.

2. Percy C, Hassoun Z, Mourad M, et al. Impact of Acute Infection Requiring Hospitalization on Tacrolimus Blood Levels in Kidney Transplant Recipients. Transplant Proc. Nov 2017;49(9):2065–2069. doi:10.1016/j.transproceed.2017.09.019

3. Wunderink HF, van der Meijden E, van der Blij-de Brouwer CS, et al. Pretransplantation Donor-Recipient Pair Seroreactivity Against BK Polyomavirus Predicts Viremia and Nephropathy After Kidney Transplantation. Am J Transplant. Jan 2017;17(1):161–172. doi:10.1111/ajt.13880

4. Brennan DC. Cytomegalovirus in renal transplantation. J Am Soc Nephrol. Apr 2001;12(4):848–55.

5. Razonable RR, Humar A, Practice ASTIDCo. Cytomegalovirus in solid organ transplantation. Am J Transplant. Mar 2013;13 Suppl 4:93–106. doi:10.1111/ajt.12103

6. Bohl DL, Brennan DC. BK virus nephropathy and kidney transplantation. Clin J Am Soc Nephrol. Jul 2007;2 Suppl 1:S36-46. doi:10.2215/CJN.00920207

7. van Rijn AL, Wunderink HF, de Brouwer CS, van der Meijden E, Rotmans JI, Feltkamp MCW. Impact of HPyV9 and TSPyV coinfection on the development of BK polyomavirus viremia and associated nephropathy after kidney transplantation. J Med Virol. Jan 9 2019;doi:10.1002/jmv.25397.

8. Rezahosseini O, Drabe CH, Sorensen SS, et al. Torque-Teno virus viral load as a potential endogenous marker of immune function in solid organ transplantation. Transplant Rev (Orlando). Jul 2019;33(3):137–144. doi:10.1016/j.trre.2019.03.004

9. Vu DL, Kaiser L. The concept of commensal viruses almost 20 years later: redefining borders in clinical virology. Clin Microbiol Infect. Oct 2017;23(10):688–690. doi:10.1016/j.cmi.2017.03.005.

10. Griffiths P. Time to consider the concept of a commensal virus? Rev Med Virol. Apr-Jun 1999;9(2):73–4.

11. Cossart Y. TTV a common virus, but pathogenic? Lancet. Jul 18 1998;352(9123):164. doi:10.1016/S0140-6736(05)77802-8

12. Walker PJ, Siddell SG, Lefkowitz EJ, et al. Changes to virus taxonomy and the International Code of Virus Classification and Nomenclature ratified by the International Committee on Taxonomy of Viruses (2019). Arch Virol. Sep 2019;164(9):2417–2429. doi:10.1007/s00705-019-04306-w

13. Haloschan M, Bettesch R, Gorzer I, Weseslindtner L, Kundi M, Puchhammer-Stockl E. TTV DNA plasma load and its association with age, gender, and HCMV IgG serostatus in healthy adults. Age (Dordr). 2014;36(5):9716. doi:10.1007/s11357-014-9716-2

14. Jaksch P, Michael K, Irene G, et al. Torque Teno Virus as a novel biomarker targeting the efficacy of immunosuppression after lung transplantation. J Infect Dis. Jul 20 2018;doi:10.1093/infdis/jiy452

15. Frye BC, Bierbaum S, Falcone V, et al. Kinetics of Torque Teno Virus-DNA Plasma Load Predict Rejection in Lung Transplant Recipients. Transplantation. Sep 17 2018;doi:10.1097/TP.0000000000002436

16. Fernandez-Ruiz M, Albert E, Gimenez E, et al. Monitoring of alphatorquevirus DNA levels for the prediction of immunosuppression-related complications after kidney transplantation. Am J Transplant. Oct 22 2018;doi:10.1111/ajt.15145

17. Beland K, Dore-Nguyen M, Gagne MJ, et al. Torque Teno virus in children who underwent orthotopic liver transplantation: new insights about a common pathogen. J Infect Dis. Jan 15 2014;209(2):247–54. doi:10.1093/infdis/jit423

18. Strassl R, Schiemann M, Doberer K, et al. Quantification of Torque Teno Virus Viremia as a Prospective Biomarker for Infectious Disease in Kidney Allograft Recipients. J Infect Dis. Sep 8 2018;218(8):1191–1199. doi:10.1093/infdis/jiy306

19. Gorzer I, Jaksch P, Strassl R, Klepetko W, Puchhammer-Stockl E. Association between plasma Torque teno virus level and chronic lung allograft dysfunction after lung transplantation. J Heart Lung Transplant. Mar 2017;36(3):366–368. doi:10.1016/j.healun.2016.10.011

20. Ruiz P, Martinez-Picola M, Santana M, et al. Torque Teno Virus is associated with the state of immune suppression early after liver transplantation. Liver Transpl. Oct 30 2018;doi:10.1002/lt.25374

21. Simonetta F, Pradier A, Masouridi-Levrat S, et al. Torque Teno Virus Load and Acute Rejection After Orthotopic Liver Transplantation. Transplantation. Jul 2017;101(7):e219–e221. doi:10.1097/TP.0000000000001723

22. De Vlaminck I, Khush KK, Strehl C, et al. Temporal response of the human virome to immunosuppression and antiviral therapy. Cell. Nov 21 2013;155(5):1178–87. doi:10.1016/j.cell.2013.10.034

23. Strassl R, Doberer K, Rasoul-Rockenschaub S, et al. Torque Teno Virus for Risk Stratification of Acute Biopsy-proven Alloreactivity in Kidney Transplant Recipients. J Infect Dis. Jan 18 2019;doi:10.1093/infdis/jiz039

24. Norden R, Magnusson J, Lundin A, et al. Quantification of Torque Teno Virus and Epstein-Barr Virus Is of Limited Value for Predicting the Net State of Immunosuppression After Lung Transplantation. Open Forum Infect Dis. Apr 2018;5(4):ofy050. doi:10.1093/ofid/ofy050

25. Takemoto AY, Okubo P, Saito PK, et al. Torque teno virus among dialysis and renal-transplant patients. Braz J Microbiol. Mar 2015;46(1):307–11. doi:10.1590/S1517-838246120131195

26. Handala L, Descamps V, Morel V, et al. No correlation between Torque Teno virus viral load and BK virus replication after kidney transplantation. J Clin Virol. Jul 2019;116:4–6. doi:10.1016/j.jcv.2019.03.018

27. Wunderink HF, Haasnoot GW, de Brouwer CS, et al. Reduced Risk of BK Polyomavirus Infection in HLA-B51-positive Kidney Transplant Recipients. Transplantation. Mar 2019;103(3):604–612. doi:10.1097/TP.0000000000002376

28. Wunderink HF, De Brouwer CS, Gard L, et al. Source and Relevance of the BK Polyomavirus Genotype for Infection After Kidney Transplantation. Open Forum Infect Dis. Mar 2019;6(3):ofz078. doi:10.1093/ofid/ofz078

29. von Elm E, Altman DG, Egger M, et al. Strengthening the Reporting of Observational Studies in Epidemiology (STROBE) statement: guidelines for reporting observational studies. BMJ. Oct 20 2007;335(7624):806–8. doi:10.1136/bmj.39335.541782.AD

30. Niesters HG. Quantitation of viral load using real-time amplification techniques. Methods. Dec 2001;25(4):419–29. doi:10.1006/meth.2001.1264

31. Maggi F, Pifferi M, Fornai C, et al. TT virus in the nasal secretions of children with acute respiratory diseases: relations to viremia and disease severity. J Virol. Feb 2003;77(4):2418–25.

32. Okamoto H, Nishizawa T, Ukita M, et al. The entire nucleotide sequence of a TT virus isolate from the United States (TUS01): comparison with reported isolates and phylogenetic analysis. Virology. Jul 5 1999;259(2):437–48. doi:10.1006/viro.1999.9769

33. Hornung R, Reed L. Estimation of Average Concentration in the Presence of Non-detectable Values. App Occup Environ Hyg. 1990;5:46–51.

34. Kalpoe JS, Kroes AC, de Jong MD, et al. Validation of clinical application of cytomegalovirus plasma DNA load measurement and definition of treatment criteria by analysis of correlation to antigen detection. J Clin Microbiol. Apr 2004;42(4):1498–504. doi:10.1128/jcm.42.4.1498-1504.2004

35. Schemper M, Smith TL. A note on quantifying follow-up in studies of failure time. Control Clin Trials. Aug 1996;17(4):343–6.

36. R: A Language and Environment for Statistical Computing. Version 3.5.2. R Foundation for Statistical Computing; 2018. https://www.R-project.org/

37. Wickham H. ggplot2: Elegant Graphics for Data Analysis. Springer-Verlag; 2016.

38. Kassambara A, Kosinski M. survminer: Drawing Survival Curves using ’ggplot2’. 2018; https://CRAN.R-project.org/package=survminer

39. Thernau T, Gramsch P. Modeling Survival Data: Extending the Cox Model. Springer; 2000.

40. Rizopoulos D. JM: An R Package for the Joint Modelling of Longitudinal and Time-to-Event Data. Journal of Statistical Software. 2010;35(9):1–33.

41. nlme: Linear and Nonlinear Mixed Effects Model. 2018.

42. van der Beek MT, Berger SP, Vossen AC, et al. Preemptive versus sequential prophylactic-preemptive treatment regimens for cytomegalovirus in renal transplantation: comparison of treatment failure and antiviral resistance. Transplantation. Feb 15 2010;89(3):320–6. doi:10.1097/TP.0b013e3181bc0301

43. Kamel M, Kadian M, Srinivas T, Taber D, Posadas Salas MA. Tacrolimus confers lower acute rejection rates and better renal allograft survival compared to cyclosporine. World J Transplant. Dec 24 2016;6(4):697–702. doi:10.5500/wjt.v6.i4.697

44. Solis M, Velay A, Gantner P, et al. Torquetenovirus viremia for early prediction of graft rejection after kidney transplantation. J Infect. Jul 2019;79(1):56–60. doi:10.1016/j.jinf.2019.05.010

45. Fernandez-Ruiz M, Albert E, Gimenez E, et al. Early kinetics of Torque Teno virus DNA load and BK polyomavirus viremia after kidney transplantation. Transpl Infect Dis. Dec 28 2019:e13240. doi:10.1111/tid.13240

46. Maggi F, Focosi D, Statzu M, et al. Early Post-Transplant Torquetenovirus Viremia Predicts Cytomegalovirus Reactivations In Solid Organ Transplant Recipients. Sci Rep. Oct 19 2018;8(1):15490. doi:10.1038/s41598-018-33909-7

47. Doberer K, Schiemann M, Strassl R, et al. Torque Teno virus for risk stratification of graft rejection and infection in kidney transplant recipients - A prospective observational trial. Am J Transplant. Feb 7 2020;doi:10.1111/ajt.15810

48. Matuschek H, Kliegl R, Vasishth S, Baayen H, Bates D. Balancing Type I error and power in linear mixed models. J Mem Lang. Jun 2017;94:305–315. doi:10.1016/j.jml.2017.01.001

49. Barr DJ, Levy R, Scheepers C, Tily HJ. Random effects structure for confirmatory hypothesis testing: Keep it maximal. J Mem Lang. Apr 2013;68(3):255–278. doi:10.1016/j.jml.2012.11.001

50. Bal A, Sarkozy C, Josset L, et al. Metagenomic Next-Generation Sequencing Reveals Individual Composition and Dynamics of Anelloviruses during Autologous Stem Cell Transplant Recipient Management. Viruses. Nov 14 2018;10(11)doi:10.3390/v10110633

