## Supplement for "Torque teno virus load predicts allograft rejection but not viral infection after kidney transplantation"

519 patients eligible KTx between 2003 and 2013 age > 18 | living donor

130 excluded - 64 no baseline sample - 65 <2 post ktx samples 2003: 11 2004: 25 2005: 30 2006: 20 2007: 13 2008: 6 2009: 4 2010: 6 2011: 6 2012: 9

389 patients included in TTV analysis

**Supplement 1** Flowchart for inclusion and exclusion of kidney transplantation recipients.

**Supplement 2** A detailed description of TTV detection

For TTV load detection DNA was extracted from 200 µL of each blood-derived sample, as described previously ^1^. To monitor the extraction efficiency and control for PCR-inhibition, irrelevant viral DNA (phocine herpesvirus) was added to the extraction and analysed by an independent PCR ^2^. DNA was eluted in a final volume of 100 μL, of which 10 μL was used as input for a TTV qPCR based on the TTV PCR primers and probe described by Maggi *et al*. ^3^ with the following oligos: sense primer, 5’-GTGCCGIAGGTGAGTTTA-3’; antisense primer, 5’-AGCCCGGCCAGTCC-3’; and sense TaqMan probe: FAM-5’-TCAAGGGGCAATTCGGGCT-3’-BHQ1. The primers target the untranslated region (UTR) of the TTV genome, which is highly conserved through the different TTV species ^4^. This means the primers target multiple TTV species at once. The qPCR reactions were performed in a total volume of 25 µl using 3.5 mM MgCl2, 0.3 µM of each primer and 0.3 µM probe. A positive PCR control was constructed by cloning the TTV target sequence from a positive patient sample into a pCR2.1-TOPO vector (Invitrogen Corporation, Carlsbad, CA, USA). This plasmid was also used to determine the qPCR TTV sensitivity and limit of detection (LOD) at 250 (2.4 log_10_) copies/ml. PCR reactions were performed by using a CFX96 real-time detection system (Biorad, Hercules, CA, USA) with the PCR cycling conditions as follow: 95°C for 15 minutes, followed by 45 cycles of 95°C for 30 seconds, 55°C for 30 seconds and 72°C for 30 seconds. Analysis of the qPCR data was performed using Bio-Rad CFX Manager version 3.1. Measured TTV loads were log_10_-transformed, as per general convention for viral load. Determined TTV loads below the LOD were set to LOD$/\sqrt{2}$ to approximate the assumed normal distribution of very weakly positive and negative loads ^5^.

**Supplement 3** A detailed description of the linear mixed effects model

A linear mixed effects model was fitted on the TTV loads. This model models the mean progression of the TTV load over time, using effects that are the same for every individual – the fixed effects – and effects that are unique for every individual – the random effects. First the random effects structure with the best fit was determined by comparing random effects structures with the likelihood ratio test using the maximum likelihood. Then the fixed effects structure was determined similarly. The highest order of the polynomial used was 3, to prevent over-fitting the model. Covariates (sex, age, underlying condition, dialysis vintage and tacrolimus vs cyclosporine as maintenance) were tested individually and added if they improved the fit of the model. The final linear mixed effect model contained a random intercept ($b_{i0}$), first and second degree terms with coefficients $b_{i1}$ and $b_{i2}$as random effects to explain the between-subject variation, and an intercept $\beta_{0}$, and terms with coefficients $\beta_{1}$, $\beta_{2}$, $\beta_{3}$, $\beta_{4}$, and $\beta_{5}$, history of dialysis and tacrolimus vs cyclosporine as fixed effects. This is written in formula as:

$y_{ij}= \beta_{0}+ \beta_{1}*t_{ij}+ \beta_{2}*t_{ij}^{2}+ \beta_{3}*t_{ij}^{3}+ \beta_{4}*{tacrolimus}_{i}+ \beta_{5}*{dialysis}_{i}+b_{i0}+b_{i1}*t_{ij}+b_{i2}*t_{ij}^{2}+\varepsilon_{ij}$

where:$t_{ij}$ and $y_{ij}$ are the timepoint and the predicted TTV load, respectively, for the *i*-th subject on the *j*-th measurement. The $\beta$-coefficients are the fixed effects that are the same for every individual, the *b*-coefficients represent the random effects unique for every patient, and the $\varepsilon_{ij}$ explains the residual variation between the observed and predicted value for each *i*-th individual and *j*-th measurement. ${tacrolimus}_{i}$ was 1 if the patient received tacrolimus, 0 if cyclosporine, and ${dialysis}_{i}$ equals 1 if the patient has a history of dialysis, 0 if otherwise.

1.	Wunderink HF, van der Meijden E, van der Blij-de Brouwer CS, et al. Pretransplantation Donor-Recipient Pair Seroreactivity Against BK Polyomavirus Predicts Viremia and Nephropathy After Kidney Transplantation. *Am J Transplant*. Jan 2017;17(1):161-172. doi:10.1111/ajt.13880

2.	Niesters HG. Quantitation of viral load using real-time amplification techniques. *Methods*. Dec 2001;25(4):419-29. doi:10.1006/meth.2001.1264

3.	Maggi F, Pifferi M, Fornai C, et al. TT virus in the nasal secretions of children with acute respiratory diseases: relations to viremia and disease severity. *J Virol*. Feb 2003;77(4):2418-25.

4.	Okamoto H, Nishizawa T, Ukita M, et al. The entire nucleotide sequence of a TT virus isolate from the United States (TUS01): comparison with reported isolates and phylogenetic analysis. *Virology*. Jul 5 1999;259(2):437-48. doi:10.1006/viro.1999.9769

5.	Hornung R, Reed L. Estimation of Average Concentration in the Presence of Non-detectable Values. *App Occup Environ Hyg*. 1990;5:46-51.
